# Network Analysis of Mental Well-being, Psychological Inflexibility, and Psychopathological Symptoms in Individuals Seeking Online Psychological Support

**DOI:** 10.64898/2026.02.22.26346851

**Authors:** Leivy Patricia González-Ramírez, Joel Omar González-Cantero, Reyna Jazmín Martínez-Arriaga, Said Jiménez, Paulina Erika Herdoiza-Arroyo, Rebeca Robles-García, Rosa Olimpia Castellanos-Vargas, Alejandro Dominguez-Rodriguez

## Abstract

**Background:** Mental well-being encompasses positive psychological functioning, life satisfaction, and engagement with daily activities. It is influenced by multiple interrelated factors, including symptoms of stress, anxiety, depression, and psychological inflexibility. Network analysis provides a data-driven framework for examining the complex interconnections between these components and for identifying elements that may play a central role in the mental well-being system. The present study aimed to identify key elements related to stress, anxiety, depression, and psychological inflexibility associated with mental well-being in individuals seeking online psychological support.

**Methods:** This cross-sectional study analyzed data drawn from the Online Well-being intervention. A total of 967 Mexican participants were included. A psychological network comprising seventy-four items was estimated, and centrality indices (strength, closeness, and betweenness) were computed to determine the relative importance of individual elements within the network. Network comparisons by gender were conducted to evaluate global and local differences.

**Results:** The network revealed multiple inter-domain associations, particularly negative relationships between mental well-being and symptoms of depression, anxiety, negative stress, and psychological inflexibility. Items reflecting self-evaluation and emotional well-being consistently emerged as the most central elements in the network across centrality metrics. Gender-based comparisons indicated overall structural similarity between networks, although differences were observed in the strength of specific connections.

**Conclusions:** Network analysis identified central elements linking mental well-being with psychological distress and inflexibility in a population seeking online psychological support. These findings contribute to a systems-level understanding of mental well-being and highlight potential targets for psychological interventions to enhance well-being and reduce distress.

## Introduction

The concept of mental well-being has evolved over time, and remains ambiguous even today, given that it may or may not include the absence of disease. It is generally agreed that it involves good physical and behavioral health, a sense of purpose in life, active participation in enjoyable work and play, pleasant relationships, and contentment (1). It is a complex construct encompassing two key psychological perspectives: hedonic, including the subjective experience of happiness and life satisfaction, and eudaimonic, comprising psychological functioning and personal fulfillment (2).

Many factors affecting individuals’ mental well-being, such as stress, anxiety, and depression, may be inversely related to, or mediate, mental well-being (3,4). Mental health problems are widespread in Mexico and rank among the top causes of disability over time (5). At the conceptual level, anxiety disorders are characterized by excessive, persistent fear and/or anxiety, heightened anticipation of threat, avoidance behaviors, and clinically significant distress or functional impairment. The Diagnostic and Statistical Manual of Mental Disorders, Fifth Edition (DSM-5) emphasizes considering cultural factors in the expression of anxiety (6). Conversely, major depressive disorder is characterized by a constellation of affective, cognitive and behavioral symptoms, including pervasive low mood, psychomotor retardation, persistent fatigue, and maladaptive cognitions such as feelings of worthlessness or excessive guilt. These features are frequently accompanied by recurrent thoughts of death or suicidal ideation and are associated with marked impairment in social, occupational, and overall functional domains (7). Finally, stress is defined as a state in which the homeostasis of the organism is challenged. It involves a combination of physiological, neuroendocrine, emotional, cognitive, and behavioral responses to stimuli or situations perceived as threatening or as exceeding an individual’s coping resources (8).

As Fonseca-Pedrero explains, (9) nosological systems, such as the DSM, conceptualize mental disorders as latent mental entities inferred from patterns of symptoms, their covariation, and duration. However, mental disorders lack clearly identifiable causes that exist independently of their symptoms, and this traditional perspective limits the understanding of their complexity. For instance, researchers have noted that anxiety, stress, and depression have been associated with various signs and symptoms. Some of these overlap with each other or are present in daily life, without being pathological per se, such as fatigue or low mood (10).

In contrast, psychological inflexibility is regarded as a transdiagnostic process linked to several psychological disorders (11–13). It is identified as a common risk factor for depression, anxiety disorders (14), and the reduction of positive emotions and sense of meaning in life (15), all of which are vital for mental well-being. However, findings have been inconsistent (16), and more studies are required across different cultures and contexts. Psychological inflexibility is defined as the rigid control of psychological responses, such as unwanted thoughts and feelings, at the expense of values-guided actions, characterized by a limited behavioral repertoire, including intolerance of ambiguity (3,12–14). Psychological inflexibility has six core aspects: experiential avoidance, inflexible attention, disrupted values, inaction or impulsivity, conceptualized self, and cognitive fusion (3).

As argued by several authors (9,17), network analysis constitutes a novel theoretical framework for the study of psychopathology. Network analysis allows interconnections between the characteristic elements assessed in mental well-being, anxiety, depression, and psychological inflexibility, to be examined and represented as nodes. The relationship between these elements is represented by edges; solid lines indicate positive associations between nodes, while dashed lines indicate negative associations. The thickness of the lines reflects the strength of the connection between nodes (11).

It has been proposed that the concept of mental health or well-being refers to a stable state within a symptom network characterized by weak interconnections among its elements. Conversely, mental disorders are associated with stable states within symptom networks displaying strong, dense interconnections. From this perspective, psychopathology not merely results from isolated symptoms, but from the dynamic interactions and mutual reinforcement among symptoms within highly connected networks (9,17). Network analysis also allows for the calculation of centrality metrics. This can help determine which nodes are more closely related to the rest of the nodes in the network, providing valuable insight into the main behaviors that may impact mental well-being (16).

Recognizing these central behaviors will enable the identification of elements that are strongly associated with mental well-being. Beyond its theoretical contribution, this framework offers practical clinical applications, guiding the identification of central symptoms for target interventions, and providing more accurate resources for the affected population (17). This approach is closer to the transdiagnostic perspective, which has become increasingly accepted in recent years, providing a new paradigm for understanding mental health, its etiology, maintenance, and clinical treatment (10).

This study seeks to identify nodes associated with stress, anxiety, and depression symptoms, as well as psychological inflexibility, which may play a key role in mental well-being using a network analysis approach in participants seeking online psychological support. The hypothesis is that mental well-being will be significantly and inversely associated with symptoms of stress, anxiety, depression, and psychological inflexibility. In addition, it will be possible to identify densely connected central nodes within the network of symptoms showing both positive and negative associations with mental well-being.

## Methods

This study was conducted and reported in accordance with the STROBE Statement (Strengthening the Reporting of Observational Studies in Epidemiology), following the checklist for cross-sectional studies. The data analyzed were drawn from the Online Well-being intervention (*Bienestar Online* in Spanish), a free, multi-country program designed to promote mental well-being in the general population. The Online Well-being intervention targets participants from seven countries: the Netherlands, Spain, Mexico, Peru, Ecuador, Brazil, and Chile (18). In Mexico, it is available at www.bienestaronline.net. To obtain a large, homogeneous sample, and to minimize potential confounding effects related to participation in the Online Well-being intervention, the present study analyzed retrospective, cross-sectional data from the subsample of Mexican participants, assessed prior to their participation in the intervention.

### Participants and Procedure

The study included nine hundred and sixty-seven participants from Mexico, recruited between December 20, 2022, and the May 14, 2024. All those who completed the pre-intervention evaluation were ≥18 years old, lived in Mexico, and gave their informed consent. Detailed information on the participants’ inclusion can be found in Figure 1.

**Figure 1.**
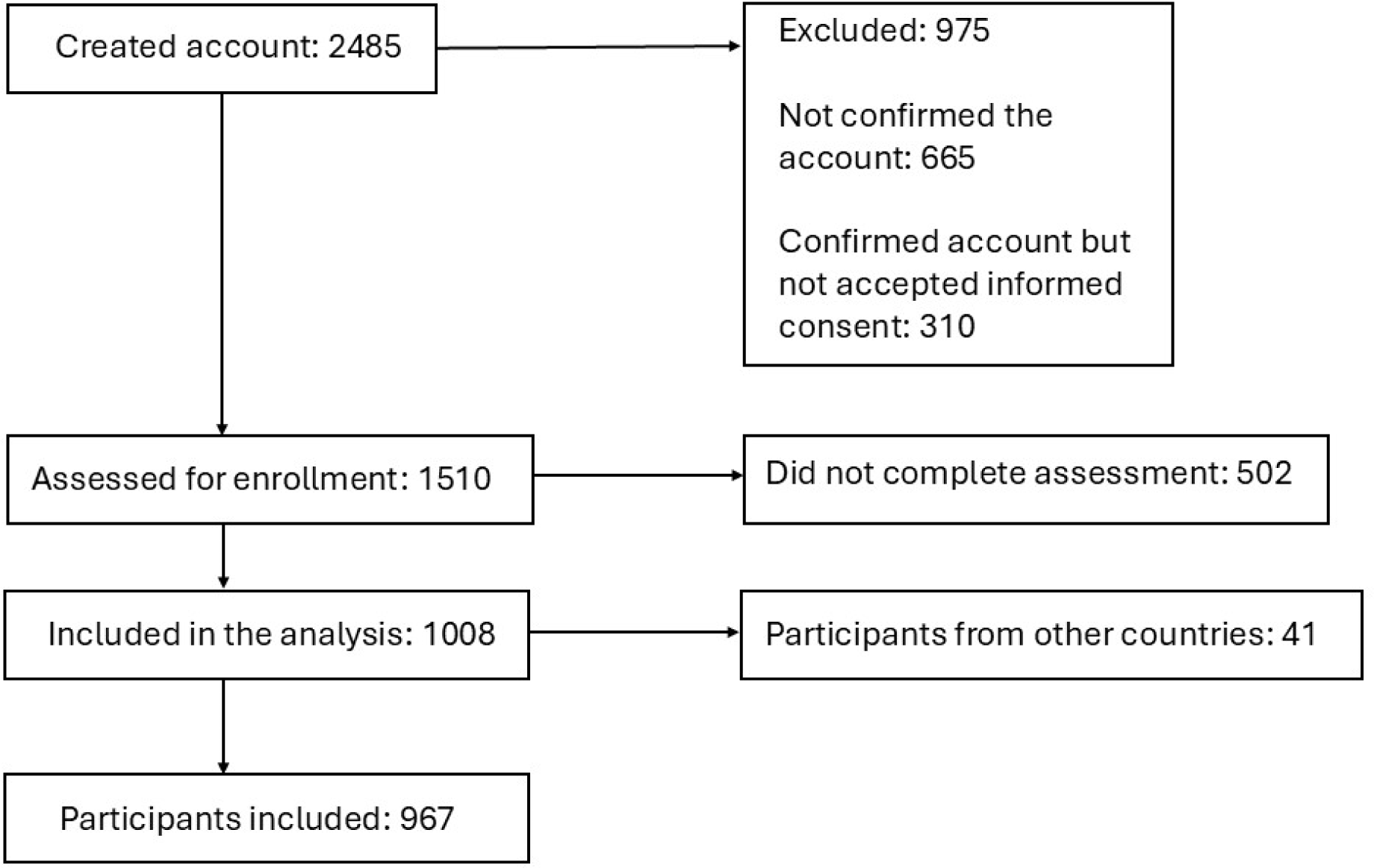
Flowchart of Study Participants

### Measures

The Warwick-Edinburgh Mental Well-Being Scale (WEMWBS) (19) is a 14-item scale measuring mental well-being based on positive aspects of mental health. Items are rated on a 5-point scale ranging from 1 (not at all) to 5 (all the time). The total score ranges from 14 to 70, with higher scores indicating greater mental well-being. The WEMWBS demonstrates strong psychometric reliability with Cronbach’s alpha scores (∝) between 0.89 and 0.91 (19). The Spanish version of the scale used in this study has demonstrated high internal consistency (∝=0.88) (20). In the current study, ∝=0.94. The items are listed in Table 1 (items 8-21).

**Table 1.** Comprehensive List of Nodes for Mental Well-being, Psychological Inflexibility, and Psychopathological Symptoms.

| Node | Item | Node | Item | Node | Item |
| --- | --- | --- | --- | --- | --- |
| 1 | Painful experiences make it hard for me to live the life I want. | 26 | I didn't sleep well. | 51 | People seemed unkind. |
| 2 | I'm afraid of my emotions. | 27 | I felt sad. | 52 | I enjoyed life. |
| 3 | I worry about not controlling my feelings. | 28 | I couldn't move forward. | 53 | I cried a lot. |
| 4 | Painful memories stop me from living fully. | 29 | Nothing made me happy. | 54 | I had fun. |
| 5 | My emotions interfere with the life I want. | 30 | I felt like a bad person. | 55 | I felt like giving up. |
| 6 | Others seem to handle life better than me. | 31 | I lost interest in my everyday activities. | 56 | I felt disliked by others. |
| 7 | My worries block my goals. | 32 | I slept more than usual. | 57 | How often have unexpected events affected you in the past month? |
| 8 | I've felt optimistic about the future. | 33 | I felt very slow. | 58 | How often have you felt unable to control important things in your life? |
| 9 | I've felt useful. | 34 | I felt restless. | 59 | How often have you felt stressed? |
| 10 | I've felt relaxed. | 35 | I wanted to die. | 60 | How often have you managed your problems? |
| 11 | I've shown interest in others. | 36 | I wanted to hurt myself. | 61 | How often have things gone well? |
| 12 | I've had extra energy. | 37 | I felt tired all the time. | 62 | How often have you felt overwhelmed by tasks? |
| 13 | I've handled problems well. | 38 | I felt unhappy with myself. | 63 | How often have you controlled your difficulties? |
| 14 | I've thought clearly. | 39 | I lost weight without trying. | 64 | How often have you felt in control? |
| 15 | I've felt good about myself. | 40 | I had trouble sleeping. | 65 | How often have you been angry about things beyond your control? |
| 16 | I've felt close to others. | 41 | It was hard to concentrate on important things. | 66 | How often have you felt that your problems were too much to handle? |
| 17 | I've felt confident. | 42 | I got irritated over small things. | 67 | Have you felt nervous or anxious? |
| 18 | I've made my own decisions. | 43 | I felt as good as others. | 68 | Have you been unable to stop worrying? |
| 19 | I've felt valued. | 44 | Everything I did felt like a struggle. | 69 | Have you worried excessively about various things? |
| 20 | I've been interested in new things. | 45 | I felt hopeful about the future. | 70 | Have you had trouble relaxing? |
| 21 | I've felt happy. | 46 | I felt like my life was a failure. | 71 | Have you felt too restless to stay still? |
| 22 | I had a poor appetite. | 47 | I felt scared. | 72 | Have you gotten irritated easily? |
| 23 | I couldn't shake my sadness. | 48 | I felt happy. | 73 | Have you felt like something terrible was about to happen? |
| 24 | I had trouble focusing. | 49 | I talked less than usual. |  |  |
| 25 | I felt depressed. | 50 | I felt lonely. |  |  |

The Center for Epidemiologic Studies Depression Scale (CESD-R) (21) is a 35-item scale designed to measure depression symptoms. Items are rated on a 5-point scale ranging from 0 (not at all or less than one day) to 4 (nearly every day for two weeks). Higher total scores are indicative of worse depressive symptoms. CESD-R demonstrates strong psychometric reliability: ∝=0.38 to 0.83. The Spanish version of the scale used in this study has demonstrated high internal consistency (∝≥0.90) (22). In the current study, ∝=0.96 (Table 1, items 22-56).

The Acceptance and Action Questionnaire II (AAQ-II) (23) is a 7-item scale measuring experiential avoidance and psychological inflexibility. Items are rated on a 7-point scale ranging from 1 (never true) to 7 (always true). Higher total scores are indicative of higher experiential avoidance and psychological inflexibility. AAQ-II has shown good psychometric properties with Cronbach’s alpha values ranging from 0.78 to 0.88. The Spanish version of the questionnaire used in this study has demonstrated adequate to high ∝ scores ranging from 0.75 to 0.93 (24). In the current study, ∝=0.92 (Table 1, items 1-7).

The Perceived Stress Scale (PSS-10) (25) is a 10-item scale measuring the stress level perceived in the previous month based on feelings and thoughts. Items are rated on a 5-point scale ranging from 0 (never) to 4 (very often). Four questions (items 4, 5, 7, and 8) are positively worded and should be reversed before adding up all the items for a total score. The higher the score, the greater the stress level. The PSS-10 has shown strong psychometric properties with a Cronbach’s alpha between 0.83 and 0.85 (26,27). The Spanish version of the scale used in this study has α = 0.83 (28). In the current study, α = 0.88 (Table 1, items 57-66).

The General Anxiety Disorder Scale-7 (GAD-7) (29) is a 7-item scale measuring the severity of generalized anxiety disorder symptoms. Items are rated on a 4-point scale ranging from 0 (not at all) to 3 (nearly every day). The maximum total score is 21. The GAD-7 screens for anxiety in individuals with no (0 to 4 points), mild (5 to 9 points), moderate (10 to 14 points), and severe symptoms (15 points or more). The Spanish versions of the GAD-7 demonstrate good reliability and validity (30). In the current study, ∝=0.89 (Table 1, items 67-73).

An online survey was used to obtain sociodemographic data including education, employment status, marital status, age, and gender.

### Ethics Approval

The ethics committee of the Universidad Autónoma de Ciudad Juárez, Mexico approved the protocol (CEI-2022-2-761), subsequently registered with ClinicalTrials.gov (NCT05443139). All participants provided their informed consent through the Online Well-being platform.

### Statistical Analyses

Individual items from the instruments were analyzed using the bootnet package in R (31) applying a Graphical Gaussian Model (GGM) with L1-regularization through graphical lasso (32). Sample size adequacy was evaluated following two widely used recommendations for cross-sectional network analysis. These include maintaining a sufficient ratio of observations to nodes (at least 5–10 participants per node) (33), and (2) assessing the accuracy and stability of the estimated network using nonparametric bootstrapping procedures (31). The analysis used 967 participants for a network of 74 nodes (N/p ≈ 13.1), surpassing recommended thresholds. All analyses were conducted with a complete case approach, using only participants who had complete data on all variables of interest.

The network was estimated using item residuals obtained after fitting linear models that adjusted for potential confounders, including age, gender, education, occupation, and marital status. The resulting network comprised 74 nodes, each representing an item, with edges reflecting the strength and direction of partial correlations between node pairs while controlling for all other variables in the network. Centrality indices—strength, closeness, and betweenness—were computed to characterize the network topology and identify the most influential nodes. Edge-weight accuracy and centrality stability were evaluated using case-dropping bootstrapping with 2,000 iterations, and stability was measured using the correlation stability (CS) coefficient. Finally, differences in network structure, global strength, and individual edges were examined by gender using the Network Comparison Test package (34). Participants who did not identify as male or female were excluded from these analyses due to the small sample size (n = 4), precluding meaningful subgroup comparisons.

## Results

Nine hundred and sixty-seven participants were included. Sociodemographic characteristics are given in Table 2.

**Table 2.** Sociodemographic Characteristics of Participants.

| Variable |  | Sex |  |  | Overall | P |
| --- | --- | --- | --- | --- | --- | --- |
|  |  | Female | Male | Other |  |  |
| n |  | 812 | 151 | 4 | 967 |  |
| Age<br>(Mean (SD)) |  | 33.85<br>(10.58) | 32.19<br>(10.62) | 28.75<br>(2.50) | 33.57<br>(10.58) | 0.13 |
| Education<br>(%) | Elementary school | 2<br>(0.24) | 0<br>(0.00) | 0<br>(0.00) | 2<br>(0.21) | 0.64 |
|  | Middle school | 13<br>(1.60) | 6<br>(3.97) | 0<br>(0.00) | 19<br>(1.96) |  |
|  | High school | 108<br>(13.30) | 20<br>(13.24) | 0<br>(0.00) | 128<br>(13.23) |  |
|  | University -<br>bachelor's degree | 493<br>(60.73) | 97<br>(64.25) | 4<br>(100) | 594<br>(61.45) |  |
|  | University -<br>master's degree | 154<br>(18.96) | 21<br>(13.91) | 0<br>(0.00) | 175<br>(18.09) |  |
|  | University -<br>Doctorate | 32<br>(3.94) | 4<br>(2.65) | 0<br>(0.00) | 36<br>(3.72) |  |
|  | Other | 10<br>(1.23) | 3<br>(1.98) | 0<br>(0.00) | 13<br>(1.34) |  |
| Work (%) | No | 263<br>(32.38) | 45<br>(29.80) | 2<br>(50.0) | 310<br>(32.06) | 0.61 |
|  | Yes | 549<br>(67.62) | 106<br>(70.20) | 2<br>(50.0) | 657<br>(67.94) |  |
| Marital<br>status (%) | Single | 452<br>(55.66) | 97<br>(64.24) | 3<br>(75.0) | 552<br>(57.09) | 0.25 |
|  | Married | 219<br>(26.98) | 37<br>(24.50) | 0<br>(0.0) | 256<br>(26.47) |  |
|  | Divorced | 64<br>(7.88) | 7<br>(4.64) | 0<br>(0.0) | 71<br>(7.34) |  |
|  | Widowed | 4<br>(0.49) | 2<br>(1.32) | 0<br>(0.0) | 6<br>(0.62) |  |
|  | Other | 73<br>(8.99) | 8<br>(5.30) | 1<br>(25.0) | 82<br>(8.48) |  |

### Network Structure

Figure 2 presents the GGM results. In this network, nodes represent individual items from mental well-being, psychological inflexibility, depression, stress, and anxiety scales. Edges correspond to estimated partial correlations between pairs of nodes, controlling for all other nodes in the network; positive associations are shown in blue, whereas negative associations are depicted in yellow. Edge thickness reflects the strength of the association. As shown in Figure 2, strong positive intra-domain relationships were observed among items assessing psychological inflexibility, depression, well-being, and anxiety. This pattern was not observed for stress-related items: instead, stress nodes separated into two distinct clusters. One comprised item reflecting negative manifestations of stress, positively associated with each other (such as nodes 57 and 58, indicating that feeling affected by unexpected events was related to feeling unable to control important aspects of life). The other comprised items indicating positive manifestations of stress, which were also positively and directly related to each other (such as nodes 61 and 64, reflecting perceptions that things are going well and feeling in control).

**Figure 2.**
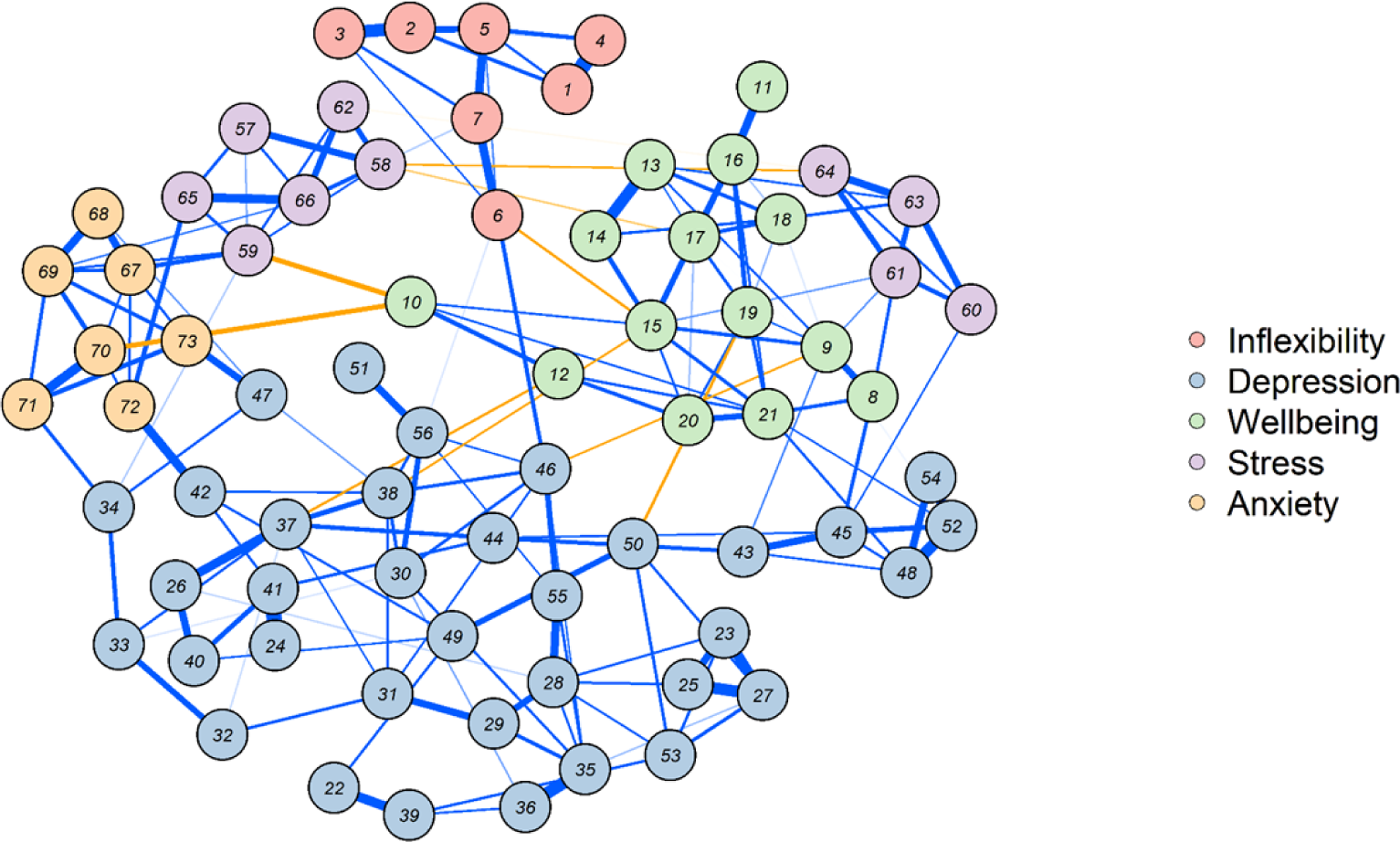
The network structure comprises individual items from psychological inflexibility, depression, well-being, stress, and anxiety scales. The Network contains 73 nodes, each representing an item, and edges indicating the level and direction of the partial correlation between pairs of nodes while controlling for the presence of the other analyzed variables. Blue edges indicate positive relationships, while yellow edges indicate negative ones. The thickness of each edge reflects the strength of the relationship between variables.

The network revealed significant inter-domain relationships. For instance, nodes representing the positive aspects of stress were positively related to well-being nodes. Conversely, nodes representing the negative aspects of stress were positively associated with anxiety nodes, which were positively related to depression nodes, which in turn were related to psychological inflexibility nodes.

The most notable inter-domain connections in Figure 2 are the negative relationships (shown in yellow) of specific nodes in the well-being domain, as they were negatively related to symptoms of depression, anxiety, negative stress, and psychological inflexibility. Examples include node 10 *(“I have felt relaxed”)*, which was strongly and negatively related to nodes 59 (feeling nervous or stressed) and 70 (difficulty relaxing). Node 15 *(“I have felt good about myself”),* was negatively related to nodes 6 *(“It seems that most people are leading their lives better than I am”)* and 38 *(“I was dissatisfied with myself”)*. This meant that the better they felt about themselves, the less they thought most people lived their lives better than them and the less dissatisfied they were with themselves. Node 9 *(“I have felt useful”)* was negatively related to node 46 *(“I thought that my life had been a failure”),* meaning that the greater the sense of usefulness, the lesser the sense of failure. Node 19 (feeling loved and valued) was negatively related to node 50 (feeling lonely) while node 12 (having plenty of energy) was indirectly related to node 37 (feeling tired all the time).

### Network Centrality

The centrality indices measured the importance of the nodes for each calculated metric, with some depression and well-being nodes consistently ranking highest across all evaluated domains (Figure 3). In terms of betweenness, nodes 38 *(“I felt unhappy with myself”)* and 15 *(“I have felt good about myself”)* scored highest, indicating that these nodes might act as key bridges or mediators in both inter- and intra-domain relationships. With regard to closeness, the highest-scoring nodes were also 38 *(“I felt unhappy with myself”)* and 15 *(“I have felt good about myself”)*. This metric identifies the nodes that communicate most efficiently with others. Finally, nodes 21 *(“I have felt happy”)* and 41 *(“Difficulty concentrating on important things”)* were the most significant in terms of strength centrality, reflecting the number and strength of the connections of these nodes.

**Figure 3.**
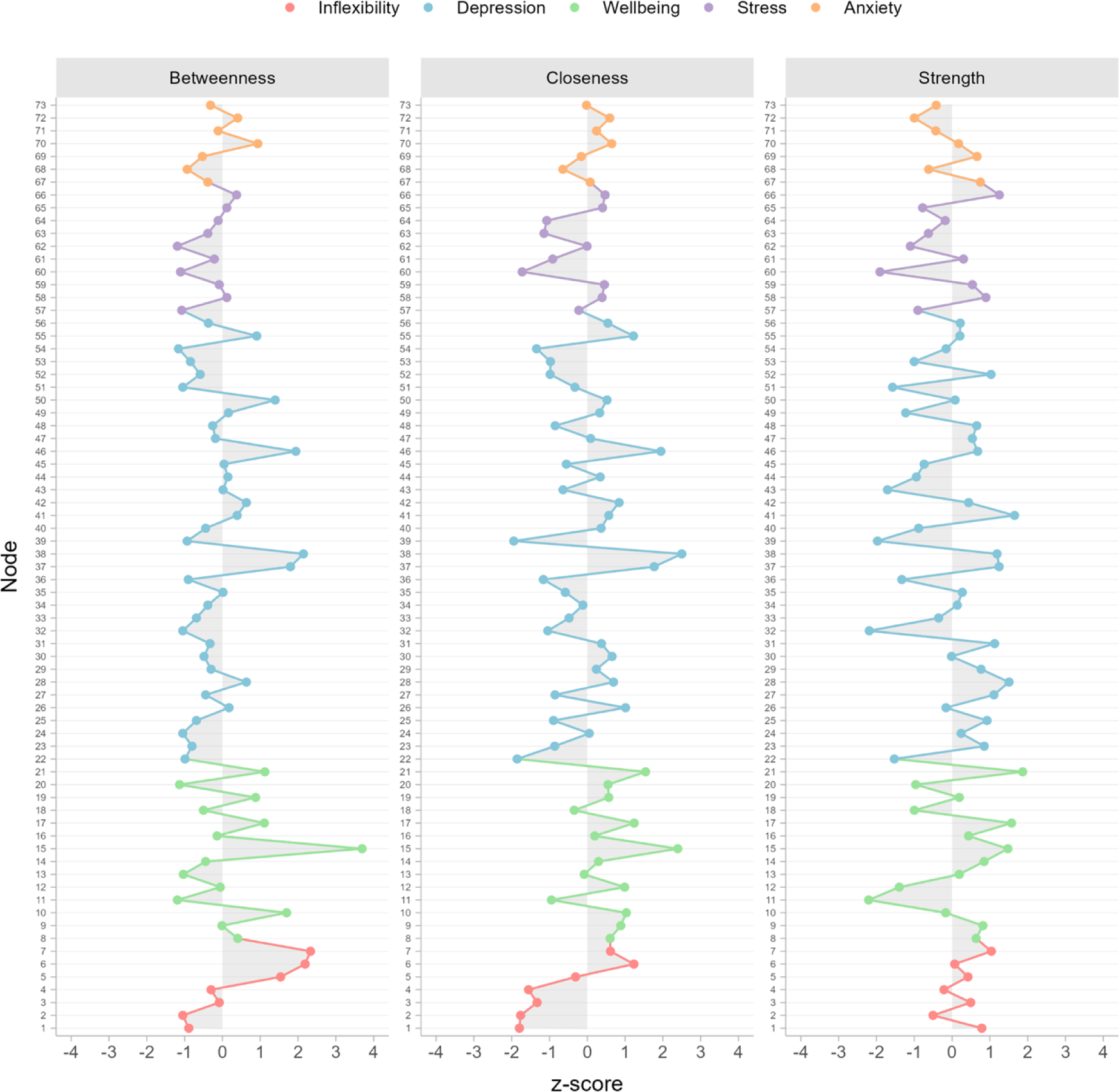
Centrality indices for each node in the network comprise individual items from the psychological inflexibility, depression, well-being, stress, and anxiety scales. The standardized Z-score, with a mean (M) = 0 and standard deviation (SD) = 1, indicates the importance of each item according to the calculated metric—the further a point shifts to the right, the greater its importance. The colors represent the various dimensions evaluated, allowing for the identification of the item with the highest intra- and inter-domain centrality.

As shown in Figure 2, node 15 *(“I have felt good about myself”)* had numerous negative relationships with nodes related to inflexibility and depression (such as nodes 15 and 6 “*Others seem to handle life better than me”*, indicating that the better people feel about themselves, the less they believe others are leading better lives). This node also had positive relationships with variables within its domain (such as nodes 15 and 17-*” I have felt confident”*-suggesting that the better people feel about themselves, the greater their self-confidence). Node 21 *(“I have felt happy”)* was strongly associated with other well-being nodes such as 20 *(“I have been interested in new things”),* 8 *(“I have felt optimistic about the future”),* 19 *(“I have felt loved and valued”),* and 15 *(“I have felt good about myself”).* The strength of this node contributes to forming a cohesive set of variables associated with overall well-being.

### Network Stability

The results for the network edges and the centrality metrics were stable, except for the betweenness index. The correlation stability (CS) coefficients exceeded the recommended 0.5 threshold. Specifically, the coefficients were 0.75, 0.75, 0.67, and 0.44 for the edges, strength, closeness, and betweenness, respectively.

### Network Comparison

The evaluation of network independence between genders using the Network Comparison Test revealed no significant differences in the Network Invariance Test (Test statistic M= 0.20, p=0.75) or the Global Strength Invariance Test (Test statistic S=0.57, p>0.40). The Edge Invariance Test showed certain statistically significant differences in the connections between the male and female networks (Figure 4). The highest number of distinct connections was found in the depression domain, with men showing more positive intra-domain relationships than women. Likewise, men displayed more intra-domain connections than women in the well-being, stress, and cognitive inflexibility domains. Regarding inter-domain connections, the link between psychological inflexibility and stress was present in men but not women.

**Figure 4.**
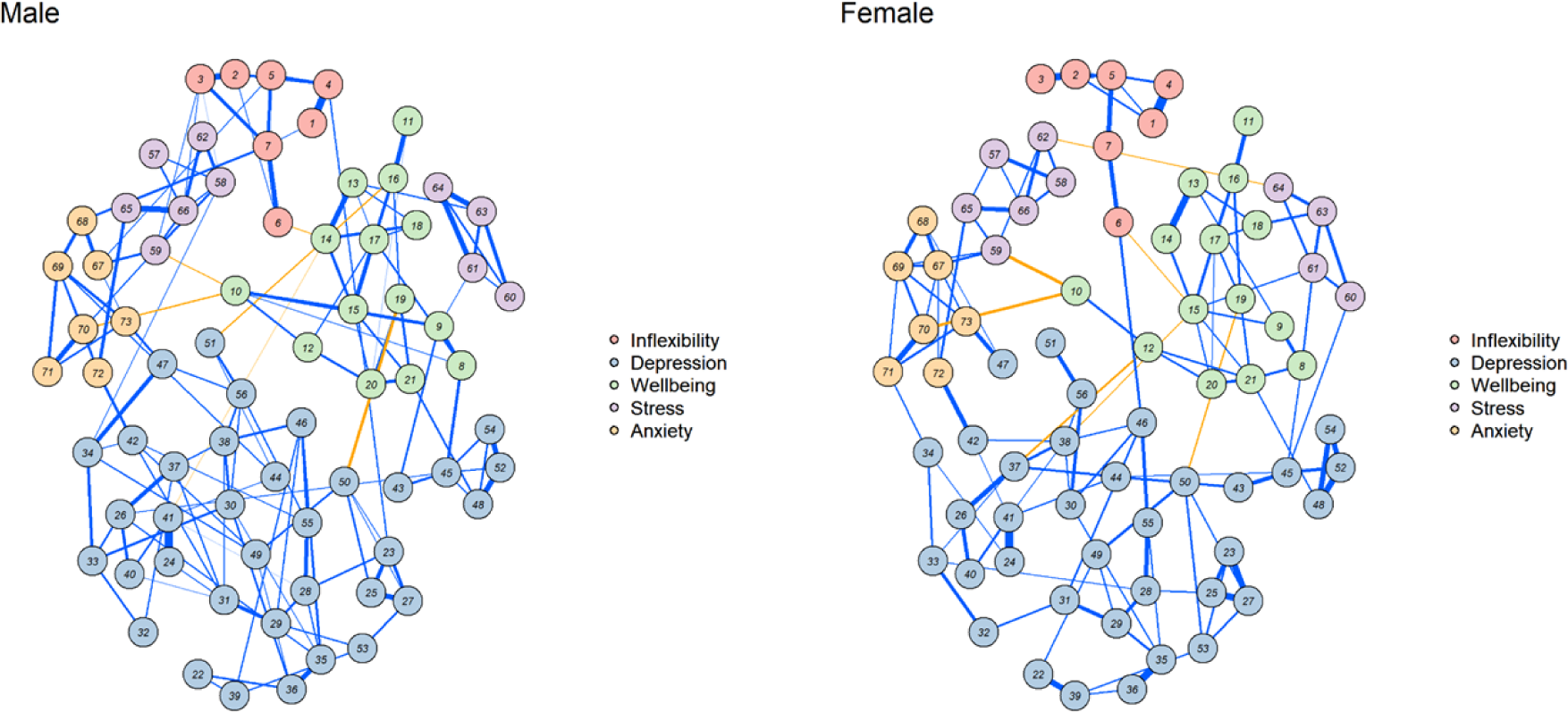
Network structures for men and women comprise individual items from the psychological inflexibility, depression, well-being, stress, and anxiety scales. This network highlights the differences found using the Network Comparison Test, showing specific connections that may be attributable to participants’ sex.

Other inter-domain connections differing between the networks are linked to specific well-being nodes. For example, the positive connection between node 18 *(“I’ve made my own decisions”)* and node 63 *(“Controlled the difficulties”)* is present in the women’s but not the men’s network. Also in women but not men, there is a negative relationship between node 12 *(“I’ve had extra energy”)* and node 37 *(“Felt tired all the time”)*, as well as between node 15 *(“Felt good about themselves”)* and both node 38 *(“Felt unhappy”)* and node 6 *(“Others seem to handle life better than me”).* These findings highlight the importance of these well-being nodes, particularly for women (Figure 4).

## Discussion

The purpose of the present study was to identify key nodes related to stress, anxiety, depression, and psychological inflexibility that may play a central role in mental well-being in a sample of individuals seeking online psychological support. As far as we know, this research is the first to use network analysis to explore how these psychological constructs are connected within this group. Network models conceptualize mental health as a system of interacting components rather than as manifestations of a single latent disorder. They are especially valuable for connecting empirical results with theory and exploring interactions between symptoms and psychological processes (17).

Overall, our findings revealed a consistent pattern in which specific well-being nodes were negatively associated with symptoms of depression, anxiety, negative stress, and psychological inflexibility. This aligns with prior research indicating that distress, depression, and anxiety are inversely related to psychological well-being, particularly when well-being is conceptualized using multidimensional frameworks such as Seligman’s PERMA model (35). Importantly, previous work has suggested that this relationship is partially mediated by psychological flexibility, a finding that is conceptually consistent with the network associations observed in the present study.

Negative aspects of stress were positively connected to anxiety nodes, which, in turn were positively associated with depressive symptoms and psychological inflexibility. For instance, the item “I have felt relaxed” showed negative associations with “feeling nervous or stressed” and “difficulty relaxing,” illustrating how affective and physiological components of stress and anxiety cluster together in the network. Psychological inflexibility, a core construct in Acceptance and Commitment Therapy (ACT), refers to rigid patterns of responding characterized by cognitive fusion and experiential avoidance (23). Conversely, psychological flexibility involves the capacity to observe internal experiences while engaging in behavior aligned with personal values (p. 11). Consistent with ACT theory, psychological inflexibility in our network was negatively related to well-being, reinforcing the notion that flexibility serves as a protective factor for mental health and flourishing, like constructs such as resilience (36,37).

The item “Others seem to handle life better than me” is a central link between psychological inflexibility and well-being. This item was directly associated with indicators of low personal satisfaction, such as “I felt like my life was a failure” and “I was dissatisfied with myself,” suggesting that social comparison may play a key role in undermining both well-being and psychological flexibility. These findings converge with evidence showing that interventions aimed at increasing psychological flexibility also lead to improvements in well-being (38). Likewise, Eadeh et al. (16) demonstrated through network analysis that well-being exerts a significant influence on ACT-related processes, further supporting the interconnected nature of these constructs.

Several well-being items emerged as particularly central in the network. Specifically, “I have felt good about myself” showed multiple negative associations with depressive symptoms and psychological inflexibility, as well as positive associations with adaptive self-evaluations. Participants reporting higher personal satisfaction were less likely to perceive others as coping better with life or experiencing self-displeasure. This finding is consistent with previous evidence indicating that low subjective well-being is a strong predictor of depressive symptoms (39).

Centrality analyses further underscored the importance of self-evaluative well-being. Nodes 38 (“I felt unhappy with myself”) and 15 (“I have felt good about myself”) exhibited the highest betweenness and closeness values, suggesting that they are central connectors between domains. Stochl et al. (40) also found that “I have felt good about myself” is a central node influencing other aspects of psychological well-being. Cross-cultural research has also demonstrated variability in key well-being items, probably due to sociocultural factors. For example, while studies in the UK and Israel identified self-regulatory clarity as central, research in India highlighted “feeling useful” as the most connected item, likely reflecting collectivist values (41).

In the Mexican sample examined here, well-being was positively associated with adaptive or positive aspects of stress, such as feeling capable of managing problems, handling challenges effectively, and maintaining a sense of control. These findings suggest that perceiving oneself as competent, useful, and capable may simultaneously enhance well-being and buffer against psychological distress. Taken together, the results emphasize self-satisfaction and perceived competence as core elements in the mental well-being network.

Gender-based network comparisons revealed invariance at the global level, indicating comparable overall structure and connectivity across men and women. However, local differences emerged at the edge level. Men exhibited stronger intra-domain associations for depression, well-being, stress, and psychological inflexibility. Conversely, women showed unique associations linking autonomy (“I’ve made my own decisions”) with perceived control over difficulties, as well as stronger connections between energy, fatigue, self-satisfaction, and social comparison. These findings suggest that although the general architecture of mental well-being may be similar across genders, specific symptom-to-symptom dynamics differ.

While denser networks and stronger edges are sometimes attributed to larger sample sizes due to their increased statistical power (34), this explanation is unlikely in the present study, as the men’s subsample (n = 151) was substantially smaller than the women’s subsample (n = 812). This points to alternative explanations, such as gender-related psychological or social differences, which should be explored in future research. Longitudinal and culturally sensitive studies are key to understanding how network dynamics change over time and across contexts.

### Implications

The theory of psychopathology posits that mental disorders are most effectively conceptualized as clusters of symptoms interconnected by causal relationships facilitating induction, explanation, prediction, and control (17). People receiving the same diagnosis can present with very different symptoms while those with different diagnoses can present similar symptoms (10). Consequently, the psychological assessment and development of interventions should not be constrained to psychopathological variables alone (42). The challenge for both mental health researchers and therapists is to determine the cause of a person’s constellation of signs and symptoms. This study analyzed specific signs and symptoms that may be relevant for both mental wellbeing and psychological distress. It identified “*I have felt good about myself*” as one of the nodes with most centrality indices-betweenness and closeness-connecting positively with mental wellbeing and negatively with stress, anxiety, depression symptoms, and psychological inflexibility. The item “*I felt unhappy with myself*” was found to be equally important. From the perspective of evidence-based psychotherapy, these data highlight the importance of exploring patient’s negative beliefs about themselves to understand and modify central factors related to depression. This converges with Beck’s cognitive theory and therapy for depression, but also reveals the relevance of positive psychology techniques designed to assess and foster patients’ recognition and appreciation of their strengths and virtues (43).

### Limitations

The present study has several limitations. First, the sample comprised individuals who agreed to participate in the Online Well-being intervention and completed the initial assessment in Mexico. They are therefore motivated to seek resources to improve their mental health and have access to technology and the Internet, which may limit the generalizability of our findings. It is also worth noting that the recruitment strategy excluded individuals without internet access and/or those lacking technological skills. Future studies could benefit from adopting more inclusive sampling procedures. Despite the novelty and potential of psychometric networks, their use is not without limitations. These include 1) sample dependency: Networks can be highly influenced by the sample collected, making it essential to implement and explicitly describe procedures to assess network stability; 2) interpretation challenges: As an exploratory, multivariate strategy, network analysis may identify relationships that are either inconsistent with theoretical frameworks or difficult to interpret. All the relationships identified in this study should therefore be subjected to further confirmation; 3) their cross-sectional nature. The networks used are based on cross-sectional data, which limits conclusions regarding the development of psychopathology and how protective nodes may help prevent mental health symptoms. Longitudinal studies could help answer these questions. Despite these limitations, it remains crucial to identify the main factors promoting mental well-being in this population, as they can inform the design of future online interventions.

## Conclusions

The findings of this article highlight key elements related to promoting mental well-being, such as the node “*I have felt good about myself,”* associated with personal satisfaction. Moreover, differences observed in intra-domain and inter-domain connections by gender raise interesting research questions about the factors that may influence the mental well-being of men and women.

## Data Availability

All relevant data are within the manuscript and its Supporting Information files.

## Acknowledgments

The authors would like to thank Joabián Alvarez Silva for the development and implementation of the Online Wellbeing platform and Mercedes Almeida, Geert van Boxtel, and Lars de Vroege from Tilburg University, the Netherlands, for their support with the project and their help in implementing the intervention in the Netherlands.

## Notes

### Competing Interest Statement

The authors have declared no competing interest.

### Funding Statement

The author(s) received no specific funding for this work.

